# Divergent amino acid and sphingolipid metabolism in patients with inherited neuro-retinal disease

**DOI:** 10.1101/2022.11.09.22281993

**Authors:** Courtney R. Green, Roberto Bonelli, Brendan R.E. Ansell, Simone Tzaridis, Michal Handzlik, Grace H. McGregor, Barbara Hart, Jennifer Trombley, Mary M. Reilly, Paul S. Bernstein, Catherine Egan, Marcus Fruttiger, Martina Wallace, Melanie Bahlo, Martin Friedlander, Christian M. Metallo, Marin L. Gantner

**Affiliations:** Molecular and Cellular Biology Laboratory, The Salk Institute for Biological Studies, La Jolla, CA, USA; Department of Bioengineering, University of California, San Diego, CA, USA; Population Health and Immunity Division, The Walter and Eliza Hall Institute of Medical Research, Parkville, VIC, Australia; Department of Medical Biology, The University of Melbourne, Parkville, VIC, Australia; Lowy Medical Research Institute, La Jolla, CA, USA; Moran Eye Center, University of Utah, Salt Lake City, UT, USA; Department of Neuromuscular Diseases, UCL Queen Square Institute of Neurology, London, UK; Medical Retina Service, Moorfields Eye Hospital NHS Foundation Trust, London, UK; University College London Institute of Ophthalmology, London, UK; University College Dublin, Dublin, Ireland

**Author notes:** These authors contributed equally to this work (Courtney R. Green, Roberto Bonelli, Brendan R.E. Ansell).

## Abstract

The non-essential amino acids serine, glycine, and alanine, as well as diverse sphingolipid species, are implicated in inherited neuro-retinal disorders and are metabolically linked by serine palmitoyltransferase (SPT), a key enzyme in membrane lipid biogenesis. To gain insights into the pathophysiological mechanisms linking these pathways to neuro-retinal diseases we performed a targeted metabolomic analysis of these pathways in sera from patients diagnosed with two metabolically intertwined diseases: macular telangiectasia type II (MacTel), hereditary sensory autonomic neuropathy type 1 (HSAN1), or both. In a cohort of >350 participants, MacTel patients exhibited broad alterations of amino acids, including changes in serine, glycine, alanine, glutamate, and branched-chain amino acids reminiscent of diabetes. MacTel patients had elevated 1-deoxysphingolipids but reduced levels of complex sphingolipids in circulation. A mouse model indicates this depletion in complex sphingolipids can be driven by low dietary serine and glycine. HSAN1 patients exhibited elevated serine, lower alanine, and a reduction in canonical ceramides and sphingomyelins compared to controls. Those patients diagnosed with both HSAN1 and MacTel showed the most significant decrease in circulating sphingomyelins. These results highlight metabolic distinctions between these two diseases, emphasize the importance of membrane lipids in the progression of MacTel, and suggest distinct therapeutic approaches.

## INTRODUCTION

Altered amino acid metabolism is increasingly appreciated as a key driver in the of pathology of a range of diseases, including metabolic syndrome, cancer, and neurological disease. Sphingolipids are critical signaling molecules and membrane components that are enriched in the nervous system and retina and synthesized from serine and fatty acyl-CoAs by serine palmitoyltransferase (SPT). When serine levels are low, alanine (or glycine) is used as a substrate by SPT to yield non-canonical 1-deoxysphingolipids that drive neuropathy and cellular dysfunction through diverse mechanisms. This highlights a potential mechanism for crosstalk between amino acid metabolism and sphingolipid biosynthesis in the context of neurological dysfunction. Numerous heritable neurological and retinal disorders are causative or linked to mutations in genes encoding sphingolipid-metabolizing enzymes, including amyotrophic lateral sclerosis (ALS), Tay-Sachs, Niemann-Pick disease, Gaucher disease, Macular telangiectasia type II (MacTel), and hereditary sensory and autonomic neuropathy type 1 (HSAN1)^1^.

MacTel is a rare (∼1:10,000) retinal degenerative disease leading to impairment of central vision and marked by reduced circulating serine and glycine levels and altered sphingolipid abundances ^2,3^. In addition to common single nucleotide polymorphisms associated with serine and amino acid metabolism, a small fraction of MacTel cases (< 3.5%) are driven by rare heterozygous coding variants in 3-phosphoglycerate dehydrogenase (*PHGDH*) or the SPT long chain subunits *SPTLC1* and *SPTLC2*^4-6^. HSAN1 is an ultra-rare (1:500,000) autosomal dominant disease caused by either SPTLC1 or SPTLC2 variants that induce preferential use of alanine over serine, promote 1-deoxysphingolipid accumulation, and drive peripheral sensorimotor neuropathy^7^. Although clinical analyses of 25 HSAN1 patients showed high penetrance of MacTel in this population, more recent studies have identified HSAN1 patients lacking macular defects^8^, suggesting the biochemical links between these diseases are more complex than first anticipated.

Altered amino acid and ceramide levels are also linked to more common diseases like type 2 diabetes (T2D), heart disease, cancer, multiple sclerosis, and Alzheimer’s disease^1,9-14^. Therefore, detailed analyses of amino acids and sphingolipid species in patients with such rare diseases may shed light on molecular driver(s) of pathogenesis and biochemical regulation relevant to MacTel, HSAN1, and more common diseases. To gain insight into the role of amino acids and ceramides in cellular health and disease, we performed a comprehensive metabolic analysis of MacTel and HSAN1 patient sera (151 Controls, 205 MacTel, and 25 HSAN1), quantifying sphingolipids, 1-deoxysphingolipids, and amino acids via targeted mass spectrometry. Our results suggests that defects in both amino acid and membrane sphingolipid homeostasis may be critical for progression of both these diseases.

## METHODS

### Human subjects

All enrolled subjects read and signed informed consent forms approved by local Institutional Review Boards, and all clinical research complied with the Declaration of Helsinki and HIPAA privacy regulations. Clinical diagnoses were made by experienced retina and neuropathy specialists and were subsequently confirmed by masked readers at the Moorfields Eye Hospital MacTel Reading Centre in London.

### Serum metabolite quantification

To investigate the levels of amino acids and sphingolipids, fasted serum was collected from MacTel and HSAN1 patients and control participants (**Table 1**). Each sample was quantified for amino acids using GC-MS and a broad panel of sphingolipids via LC-MS/MS (**Supplemental Table 1**).

**Table 1:**
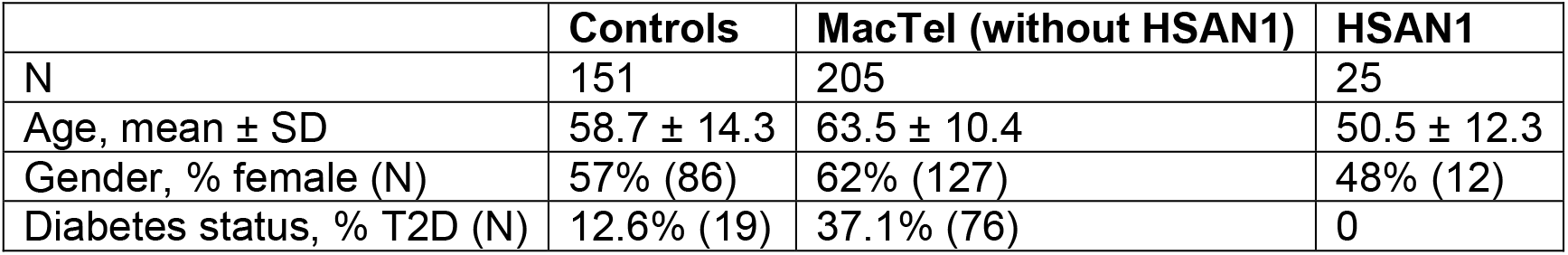
Study cohort composition statistics.

### Amino acid GC-MS analysis

In parallel to the SL extraction, 10µL of patient or control serum was aliquoted and extracted as previously described^15^. Briefly, 10µL of diluted Cambridge Isotopes I.S. mix (MSK-A2-1.2) was added, followed by 80µL of methanol. Samples were vortexed for 30 seconds, then centrifuged at 21,000g for 10 minutes at 4°C. 80µL of supernatant was collected and evaporated to dryness in a 4°C centrivap. Dried polar metabolites were derivatized in 2% (w/v) methoxyamine hydrochloride in pyridine and incubated at 45°C for 60 min. Samples were then silylated with N-tertbutyldimethylsilyl-N-methyltrifluoroacetamide (MTBSTFA) with 1% tert-butyldimethylchlorosilane (tBDMCS) (Regis Technologies) at 45°C for 30 min. Polar derivatives were analyzed by GC-MS using a DB-35MS column (30m x 0.25mm i.d. x 0.25μm, Agilent J&W Scientific) installed in an Agilent 7890B gas chromatograph (GC) interfaced with an Agilent 5977A MS with an XTR ion source. The MS source was held at 230°C and the quadrupole at 150°C with helium as the carrier gas. The GC oven was held at 100°C for 2 min, increased to 300°C at 10°C/min, held at 300°C for 4 min, then held at 325°C for 3 min.

### Sphingolipid LC-MS/MS analysis

Human sera were randomly assigned across eight batches for extraction. Each batch contained 50 patient samples and 11 quality control serum samples. Batches were not significantly different in terms of patient age or sex.

Samples were thawed in a 4°C cold room and vortexed quickly to remove gradients. 100µL of patient or control serum was aliquoted, and 20µL of I.S. mix (**Supplemental Table 1**) was added, followed by 980µL of 1:1 1-Butanol:Methanol (BUME). Samples were vortexed for 2 minutes in a cold room, then sonicated for 30 minutes in a cold room. Samples were then centrifuged at 4°C for 5 min at 14,000g. Supernatant was dried under nitrogen gas and immediately resuspended in 100µL buffer containing 100% methanol, 1 mM ammonium formate and 0.2% formic acid (Buffer B) followed by a quick vortex, 5 minute sonication, and 5 minute 4°C centrifugation at 14,000g. Supernatant was transferred to a fused glass vial and loaded onto the LC-MS/MS.

SL species were determined via liquid chromatography mass-spectrometry (Agilent 6460 QQQ). SLs were separated on a C8 column (Spectra 3 µm C8SR 150 × 3mm ID, Peeke Scientific, CA) utilizing transitions from Burla et al^16^ and quality control described in Broadhurst et al^17^. Mobile phase A was composed of 100% HPLC grade water containing 2 mM ammonium formate and 0.2% formic acid and mobile phase B consisted of 100% methanol containing 0.2% formic acid and 1 mM ammonium formate. The gradient elution program held at 82% B for 3 min, raised to 90% B over 1 min and linearly increased to 99% B over 14 min, maintained for 7 min, and returned to 82% B over 2 min. The 82% mobile phase B was maintained for 3 min followed by re-equilibration for 10 min. The capillary voltage was set to 3.5 kV, the drying gas temperature was 350 °C, the drying gas flow rate was 10 L/min, and the nebulizer pressure was 60 psi.

A new C8 column was installed immediately prior to Batch 1 and was used for all eight batches. Prior to each batch, fresh buffer was made, the source was cleaned, and 100% acetonitrile was flushed through the column for 30-45 minutes. The column was equilibrated with 14 total injections of quality control samples, a procedure blank, and internal standard mixture^17^. Quality control samples were injected throughout the sequence to identify any machine drift. Blank injections were never performed to minimize variation of sample matrix in the column. SL species were analyzed by SRM of the transition from precursor to product ions at associated optimized collision energies and fragmentor voltages (**Supplemental Table 1**). SLs were then normalized to an internal standard of known concentration followed by batch correction and further statistical analyses.

### Mouse husbandry, diet, and metabolite quantification

All animal procedures were approved by The Scripps Research Institute Animal Care Committee and we adhered to all federal animal experimental guidelines. At eight weeks of age, male C57/Bl6J mice were fed amino acid adjusted diets for 3-5 weeks as indicated. Mice were fed ad libitum isocaloric and isonitrogenous control or serine- and glycine-free custom diets (Envigo Teklad Diets, Madison, WI) (**Supplemental Table 2**). Blood was collected at 6 AM, and serum was isolated after centrifugation at 2,000g.

As described above for the human serum, 10µl of mouse serum was extracted for amino acid quantification via GCMS and 50µl of serum was extracted for sphingolipid quantification using LCMS. All samples were run in a single batch and quantified using internal standards described.

Mouse serum was extracted using 500µL MeOH, 400µL saline, 100µL of water, and 19µL of internal standard mix containing C13-dihydroceramide-d7, C15-ceramide-d7, d18:1-d7/15:0 glucosylceramide, d18:1-d7/15:0 lactosylceramide, and d18:1/18:1-d9 sphingomyelin. One mL of - 20°C chloroform was added. Samples were vortex mixed for 5 min and spun down for 5 min at 4°C at 15,000 g. The organic phase was collected and 2μL of formic acid was added to the remaining polar phase which was re-extracted with 1mL of -20°C chloroform. Combined organic phases were dried and the pellet was resuspended in 100μL of buffer containing 100% methanol with 1 mM ammonium formate and 0.2% formic acid.

### Imaging quantification

Imaging data sets included color fundus photographs (CFP), optical coherence tomography scans (OCT; volume scans of 15° × 10° [high-resolution mode, 97 scans] and 30° × 25° [high-speed mode, 61 scans]; Spectralis; Heidelberg Engineering, Heidelberg, Germany), fundus fluorescein angiograms (FFA), blue wavelength autofluorescence images (BAF), and blue-light reflectance images (BLR; all HRA Spectralis; Heidelberg Engineering, Heidelberg, Germany). Macular pigment optical density (MPOD) was measured using dual-wavelength autofluorescence (HRA Spectralis; Heidelberg Engineering, Heidelberg, Germany). Only complete, high quality image datasets were considered for the final analysis. CFP were graded for the presence of retinal crystals, pigment plaques, and neovascular membranes. The presence of retinal crystals was additionally verified on blue-light reflectance images as previously described^18^. OCT volume scans were analyzed for the presence of ellipsoid zone (EZ)-loss, outer retinal hyper-reflective changes, subretinal/sub-RPE neovascular membranes, and neovascular exudation. Eyes were classified into stages 0-4 using a simplified OCT-based disease staging system that was modified according to the classification proposed by Chew et al^19^. Stage 0 was defined as no visible break of the EZ, no outer retinal hyper-reflectivity (ORHR), and no neovascularization; stage 1 as break of the EZ but absence of ORHR; stage 2 as break of the EZ, presence of ORHR, but absence of subretinal/sub-RPE neovascular membranes; stage 3 as break of the EZ, presence or absence of ORHR, presence of neovascular membranes without detectable neovascular activity. Stage 4 was defined as break of the EZ, presence or absence of ORHR, and presence of exudative neovascular membranes and/or fibrotic neovascular scars. Additionally, eyes were graded for hyper-reflectivity, and classified into three different categories (classes 0-2). Class 0 was defined as lack of ORHR and neovascular membranes; class 1 as presence of ORHR, but absence of neovascular membranes; and class 2 as presence of neovascular membranes. Definitions for ORHR, subretinal/sub-RPE neovascularization and neovascular exudation are in accordance with previously published descriptions^20^.

For quantification of EZ-loss, OCT volume scans were analyzed using the 3D view panel as previously described^21^. MPOD maps were analyzed and eyes were assigned to MPOD-classes 1 to 3 according to the classification proposed by Zeimer et al^22^. In short, this classification differentiates between three distinct distribution patterns (“MPOD-classes”) of macular pigment, and outlines a depletion of macular pigment that increases with MPOD-classes.

## Statistical Analysis

To estimate machine measurement drift (analytical variation) over time in each machine-batch-metabolite group, mass spectral abundances were log-transformed, and a third-order polynomial equation was fitted using all experimental samples. As a small proportion of metabolite measurements were missing across the dataset (average 1.2%, SD 4.1%), we performed imputation using the GSimp R package^23^ under a ‘missing not at random’ assumption—that is, the missing metabolite abundances were assumed to fall near the lower limit of detection. Comparison of the imputation results for the total data, compared with a subset excluding HSAN1 patients, indicated negligible differences (**<u>Supplemental F</u>**igure 1A). Using the lme4 package^24^, a mixed effects linear model was fitted to test the association between metabolite abundance and disease status correcting for age, sex, T2D status, batch and machine drift (fixed effects), with a random intercept for groups of related individuals. The three disease groups were first compared with healthy controls. Subjects with MacTel only, or MacTel and HSAN1, were then compared to HSAN-only subjects. Each of the five contrasts were independently corrected for false discovery rate using the Benjamini Hochberg method. The partial residuals from subjects with any of the three diseases were calculated for plotting and for metabolic co-abundance analysis. The corrplot package was used to construct disease-specific metabolite co-abundance plots, and the R tidyverse suite of packages^25^ were used for general data transformation, residualization and visualization.

Mouse metabolic levels where log2 transformed and scaled to have zero mean and one standard deviation. Association between mouse metabolic levels and diet was assessed via linear regression modeling. Association models were performed separately for amino acids and lipids. Correction for false discovery rate was performed using the Benjamini Hochberg method.

## RESULTS

### MacTel patients exhibit altered levels of non-essential amino acids and sphingolipids

To investigate amino acid and sphingolipid changes in MacTel patients, we compared metabolite abundance in fasted serum from 205 MacTel patients and 151 healthy controls (**Table 1; Supplemental Table 3**). Differences in age, gender, and prevalence of diabetes were corrected for when testing for association. Consistent with prior results, MacTel patients had significantly lower levels of circulating serine and glycine compared to controls (**Figure 1A**). More than 75% of the MacTel patients had lower glycine levels than the mean level for control participants (**Supplemental Figure 1B**). MacTel patients also had higher circulating levels of alanine, glutamate, and the branched-chain amino acids (BCAAs) leucine and valine, suggesting that amino acid dysregulation in these patients is similar to that observed in diabetic patients^26^ (**Figure 1A**).

**Figure 1.**
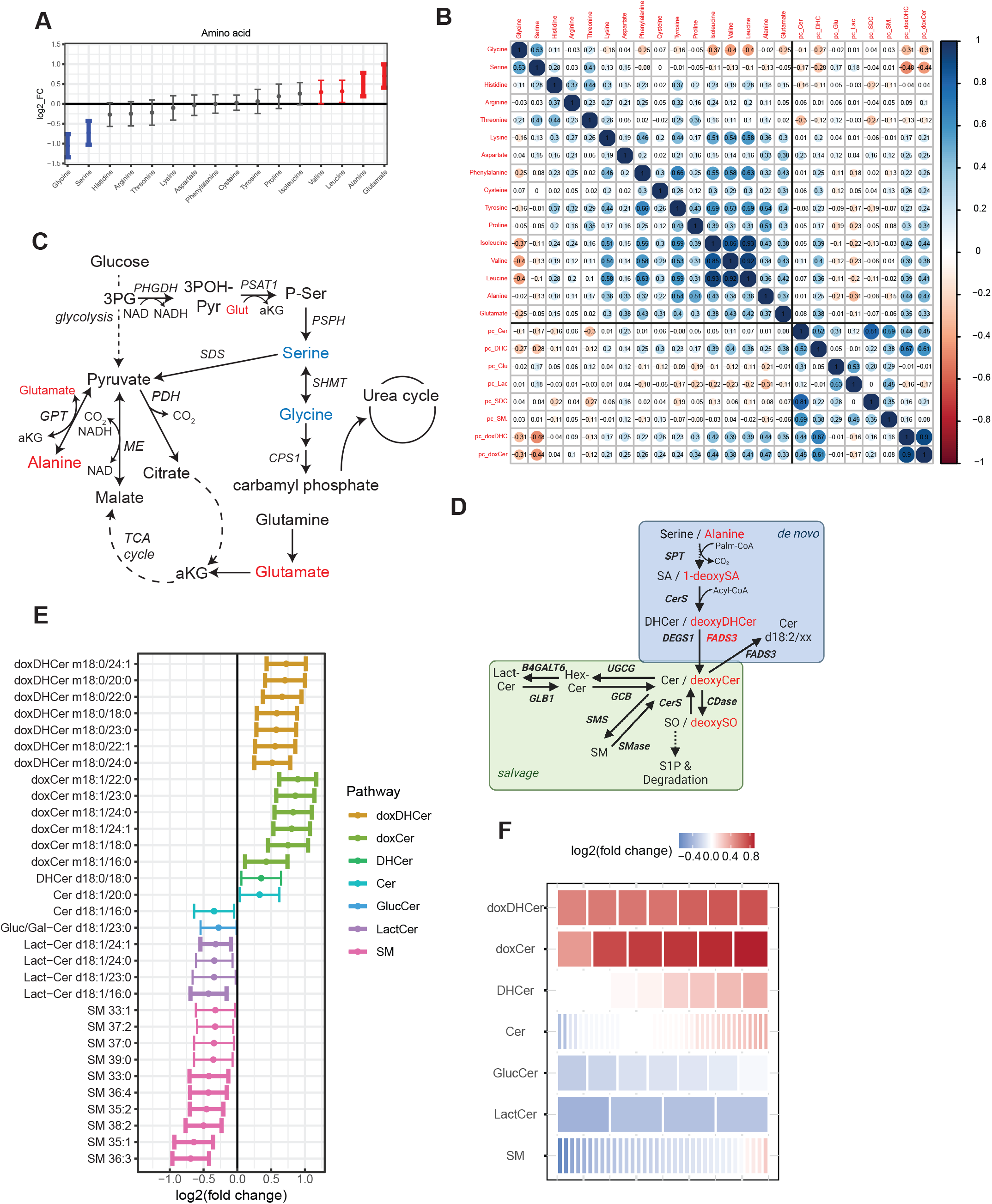
MacTel patients exhibit altered levels of non-essential amino acids and sphingolipids A. Log-fold change of nominally significant metabolites in 16 amino acids in MacTel subjects compared to Controls. Points denote the log-fold change and bars extend to the 95% confidence interval. Bold bars indicate significance after correction for FDR. B. Correlation table of amino acids and lipids principal components in Controls. Numbers indicate Pearson’s correlation coefficient (R). Abbreviations in this panel: Cer, ceramide; DHC, dihydroceramide; Glu, glucosyl-ceramide; Lac, lactosyl-ceramide; SDC, ceramide (d18:2/xx); SM, sphingomyelin; doxDHC, deoxydihydroceramide; doxCer, deoxyceramide C. Simplified metabolic map demonstrating the connections between glycolysis, TCA cycle, amino acid metabolism, and the urea cycle. D. Map of SL metabolism with the *de novo* and salvage pathways indicated. E. Change in all measured sphingolipids (SL) between MacTel subjects compared to Controls, grouped by SL class. The tiles in each row represent lipids contained in that class, colored by log fold change in MacTel compared to Controls. The color blue represents depletion and red represents increased abundance. F. Changes in abundance across SL classes. Each row is composed of tiles representing lipids contained in the group. The color of each tile represents the mean log fold change of that lipid in MacTel patients to Controls. The color blue represents depletion and red represents increased abundance. Abbreviations: doxDHCer, deoxydihydroceramide; doxCer, deoxyceramide; DHCer, dihydroceramide; Cer, ceramide; Gluc/Gal-Cer or GlucCer, glucosyl/galactosyl-ceramide; Lact-Cer, lactosyl-ceramide; SM, sphingomyelin

We also analyzed pairwise correlations within MacTel and control groups between all 16 measured amino acids (**Figure 1B & Supplemental Figure 1C**). Interestingly, alanine and glutamate correlated positively to each other (as well as with other essential and non-essential amino acids) in control participants, and this association was diminished in MacTel patients (**Figure 1B & Supplemental Figure 1C**). A similar trend was also noted with glycine, which correlated negatively with BCAAs in control participants but exhibited a weaker association in MacTel patients (**Figure 1B & Supplemental Figure 1C**). Many of these associations are expected and arise due to transaminase activity in the liver or kidney, which are hubs for nitrogen metabolism that exchange nitrogen between glutamate and specific amino acids. The weaker correlations in MacTel patients could be indicative of diminished nitrogen homeostasis, as CPS1, an enzyme participating in the urea cycle, has also been identified as having a protective effect in MacTel patients^2^ (**Figure 1C)**.

SPT synthesizes sphingoid bases by condensing serine and fatty acyl-CoAs but can also use alanine as a substrate when mass action favors this reaction, producing 1-deoxysphingoid bases. These can be metabolized to deoxy(dihydro)ceramides but not further along the canonical sphingolipid pathway (**Figure 1D**). We previously demonstrated that 1-deoxysphinganine (m18:0) from hydrolyzed serum sphingolipids was elevated in MacTel patients and correlated strongly with serine and alanine levels^4^. To assess how intact canonical and non-canonical sphingolipids were altered we performed a targeted analysis of species using LC-MS/MS (**Figure 1E-F**). Normalized abundances of all lipid species based on class-specific internal standards were determined in all patient groups (**Supplemental Figure 2A-D, Supplemental Table 4**). As expected, all detected deoxydihydroceramides and deoxyceramides were significantly elevated in MacTel patients, and these species were 35-85% higher compared to control subjects, correlating strongly with serine and alanine (**Figure 1E-F, Supplemental Figure 2A and E**). We quantified eight dihydroceramide species and 20 ceramides (13 with 18:1 bases and seven with 18:2 bases). Despite having reduced serine in circulation, dihydroceramides and ceramides (d18:1 and d18:2) were not broadly decreased in MacTel patients (**Figure 1E-F, Supplemental Figure 2B**). In contrast to 1-deoxysphingolipids, canonical ceramides are further metabolized in the Golgi to form glycosphingolipids and sphingomyelins. We quantified seven glucosyl/galactosyl-ceramides (Gluc/Gal-Cer), four lactosylceramides (Lact-Cer) and 35 sphingomyelin (SM) species, and many (15/46) were significantly decreased in MacTel patients compared to control participants (**Figure 1D-F**). In particular, glycosphingolipids of relatively high abundance such as Lact-Cer d18:1/24:1, Lact-Cer d18:1/24:0, and Lact-Cer d18:1/16:0 were significantly decreased in MacTel patients (**Figure 1E-F and Supplemental Figure 2C**). On the other hand, lower abundance but distinct (odd-chain or unsaturated) SM species were reduced nominally in MacTel patients (**Figure 1E-F and Supplemental Figure 2D**). The decrease in complex sphingolipids without concomitant decreases in the dihydroceramides may indicate an SPT-independent mechanism is driving these reductions.

To better understand how these circulating amino acids and sphingolipids might be co-regulated we performed a correlation analysis between each amino acid and the principal components derived from each sphingolipid class. Correlations for control subjects, MacTel patients, and the difference between them are depicted in **Figure 1B & Supplemental Figure 1C**. Serine, glycine, alanine, and to a lesser extent the BCAAs all strongly correlated with deoxysphingolipid abundances. On the other hand, canonical sphingolipids showed minimal correlation with amino acids aside from dihydroceramides, which showed a positive relationship with BCAAs and a negative correlation with serine. Sphingolipid classes correlated strongly with one another (>0.5) when they were separated by a single reaction (i.e dihydroceramide and ceramide or Gluc/Gal-Cer and Lact-Cer). These results may reflect the flux topology of sphingolipid metabolic networks, where synthesis and recycling of complex sphingolipids occurs at higher rates than sphingoid synthesis/turnover (and the lack thereof for 1-deoxysphingolipids). However, they may also suggest that amino acid dysregulation in MacTel patients may be unrelated to alterations in canonical membrane sphingolipids.

### Low serine and glycine levels reduce circulating ceramides and sphingolipids in mice

The lack of correlation between serine and the canonical sphingolipids might suggest that the decrease in more complex sphingolipids such as Gluc/Gal-Cer and Lact-Cer observed in MacTel is a serine-independent change. To test if reduced levels of serine and glycine (as observed in MacTel) can cause changes in canonical and complex sphingolipid levels, we fed mice amino acid altered diets, either control or a diet lacking serine and glycine but with increased levels of other non-essential amino acids to make them isonitrogenous and isocaloric (**Figure 2A, Supplemental Table 2**). Mice fed the serine/glycine deficient diet had 58% and 75% reduction in circulating serine and glycine levels respectively compared to mice fed the control diet and no other amino acids were significantly altered (**Figure 2B**). As expected, the mice fed the serine/glycine deficient diet had elevated serum levels of deoxydihydroceramides and deoxyceramides compared to control mice. The reduction in serine and glycine also led to a broad decrease in the canonical and complex sphingolipids (**Figure 2B, Supplemental Fig 3A-D**). Distinct from the MacTel patients, dihydroceramides showed a trend to be reduced and almost all ceramides were nominally or significantly decreased (**Figure 2B**). The complex sphingolipids also were broadly reduced, with all four Gluc/Gal-Cer species significantly reduced and a trend for Lact-Cer and SM to be decreased.

**Figure 2.**
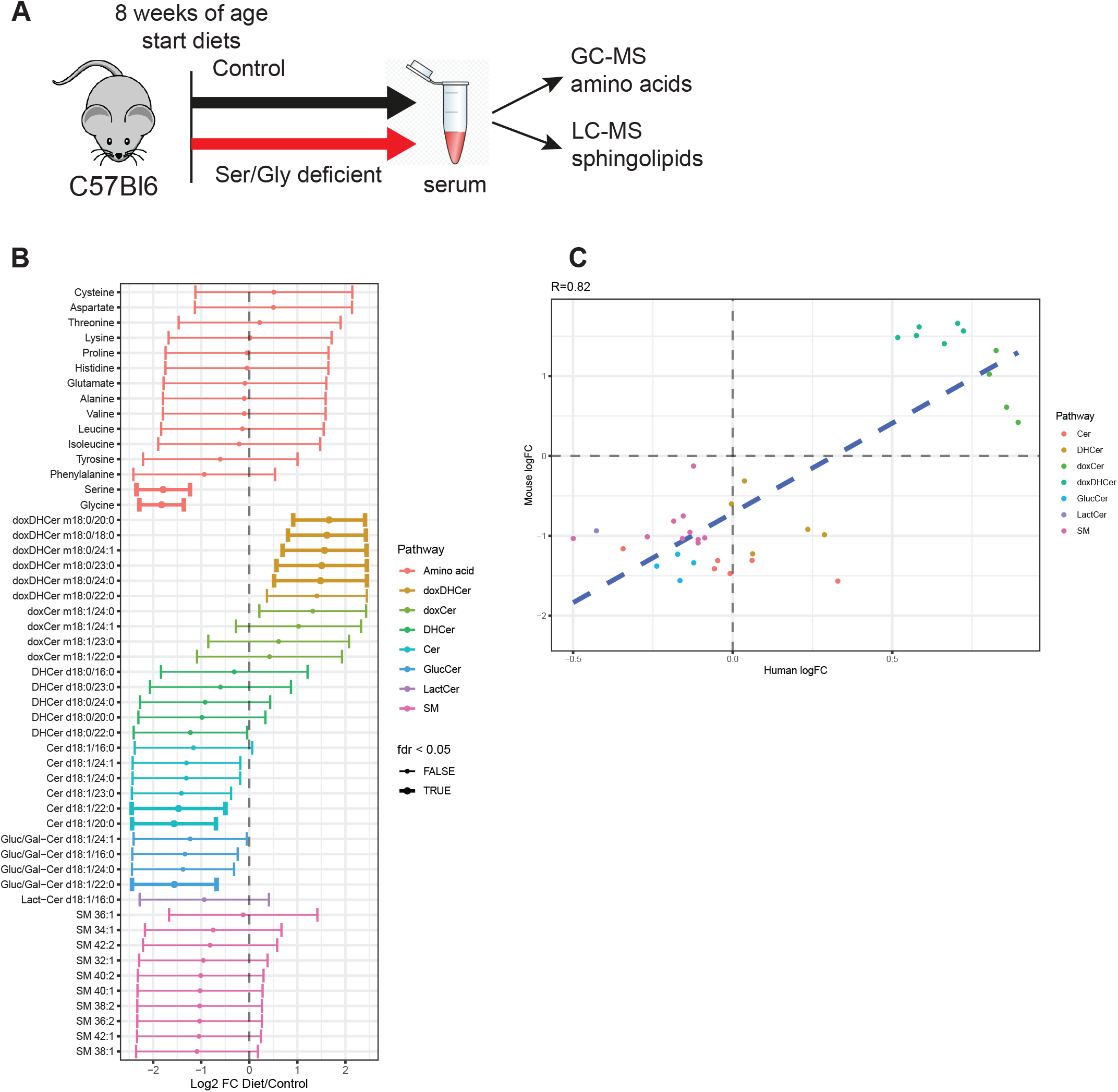
Low serine and glycine levels reduce circulating ceramides and complex SL in mice A. Log-fold change of all tested metabolites in diet mice subjects compared to control diet mice. Points denote the log-fold change and bars extend to the 95% confidence interval. Bold bars indicate significance after correction for FDR. B. Correlation plot between human and mice Log-fold change. Pearson correlation coefficient is reported at the top of the figure and the correlation line is displayed in blue. Amino-acids are not contained in this figure as they were artificially altered by design in the mice experiment. Abbreviations: doxDHCer, deoxydihydroceramide; doxCer, deoxyceramide; DHCer, dihydroceramide; Cer, ceramide; Gluc/Gal-Cer or GlucCer, glucosyl/galactosyl-ceramide; Lact-Cer, lactosyl-ceramide; SM, sphingomyelin

We compared sphingolipid changes in the mice driven by low serine and glycine to those of the MacTel patients and found strong agreement (Pearson’s R=0.82) (**Figure 2C**). Although a reduction in simple ceramides in glycine/serine-deprived mice is not recapitulated in MacTel subjects, these results indicate the reduction in complex sphingolipids (Gluc/Gal-Cer, Lact-Cer and SM) observed in MacTel can be induced by limiting serine and glycine availability.

### MacTel patients presenting neo-vascularization have reduced levels of sphingomyelins

To explore potential associations between metabolite levels and MacTel patient disease status, we evaluated retinal imaging markers known to be associated with disease severity^20-22,27-29^, analyzing multimodal retinal images of 200 eyes from 100 MacTel patients within this cohort (**Figure 3A**). These patients were selected at random and exhibited no significant differences in measured metabolite levels relative to the non-selected patients. Age of disease onset and rate of disease progression were not available for this cohort. We subsequently compared the graded phenotypic values to patient metabolite levels. Individual metabolites and principal components derived from lipid classes were tested for retinal phenotype associations. Both individual eye values (as well as their sum) were tested (**Supplemental Tables 5-6**). Age of disease onset and the rate of disease progression were not available for this analysis. We found that SM negatively correlated with neovascularization, a phenotype frequently observed in MacTel and indicative of late-stage disease (**Figure 3B-D**)^20,28,30,31^. Additionally, hyper-reflectivity and disease stage were also found to be negatively correlated with SM levels, although this association was predominantly driven by their relationship with neovascularization (**Supplemental Figure 4)**.

**Figure 3:**
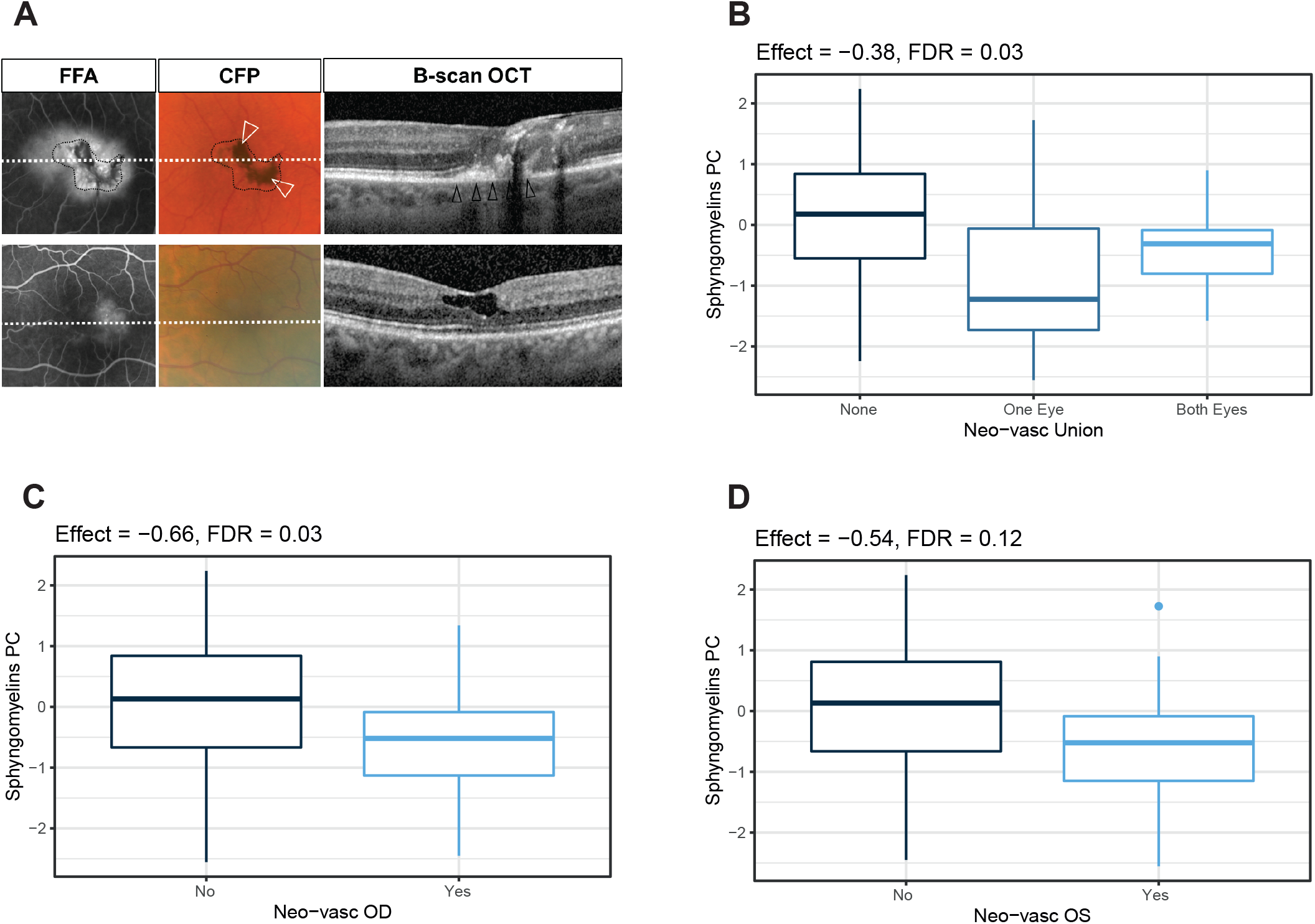
MacTel patients presenting neovascularization have reduced levels of sphingomyelins. A. Representative left eyes of two MacTel patients with low (top) and high (bottom) sphingomyelin serum levels. Eye with a large subretinal/sub-RPE neovascular membrane (black arrowheads) and pigment accumulation (white arrowheads) (top) and eye with mild parafoveal leakage on fundus fluorescein angiography (FFA) and lack of hyper-reflective changes on OCT (bottom). B. Association between sphingomyelins principal component score and neovascularization in both eyes (left + right). Center line indicates the median and box boundaries denote the 25th and 75th percentiles. C. Association between sphingomyelins principal component score and neovascularization in right eyes (OD) only. Center line indicates the median and box boundaries denote the 25th and 75th percentiles. D. Association between sphingomyelins principal component score and neovascularization in left eyes (OS) only. Center line indicates the median and box boundaries denote the 25th and 75th percentiles.

### HSAN1 patients have altered circulating levels of canonical ceramides and amino acids

Given the links between MacTel, HSAN1, and doxSLs, we analyzed serum from 25 HSAN1 patients carrying six different variants in either *SPTLC1* or *SPTLC2* (**Table 2 and Supplemental Table 7**). Patients with the *SPTLC1* C133Y or *SPTLC2* S384F variants have both peripheral neuropathy and MacTel. Patients with the remaining four variants studied here, while having severe peripheral neuropathy, do not show signs of retinopathy (**Table 2 and Supplemental Table 7**)^8^. While several patients carrying the *SPTLC1* C133Y variant and one with the C133W variant were taking serine to treat their disease at the time of serum collection (**Supplemental Table 7**), we compared effect sizes from analyses with, and without these patients and found that exclusion of these patients had little effect on the overall group statistics other than elevating serine itself (R=0.94, p<1E-10) (**Supplemental Figure 5**). Comparing HSAN1 subjects (independent of their MacTel status) to control participants, neuropathy patients exhibited high abundances of deoxydihydroceramides and deoxyceramides with a variety of acyl chains lengths (**Figure 4A-B, Supplemental Figure 6A**). Interestingly, the canonical ceramides were broadly reduced in HSAN1 patient sera compared to controls, including dihydroceramides, d18:1 and, most strikingly, d18:2 ceramides (**Figure 4A-B, Supplemental Figure 6B**). While SMs were also broadly decreased, the other complex sphingolipids, Gluc/Gal-Cer and Lact-Cers, were not uniformly altered (**Figure 4A-B, Supplemental Figure 6C-D**). While the numbers of patients with individual variants are low, these changes were uniform across variants (**Supplemental Figure 7A-G**). These findings suggest HSAN1 patients have decreased canonical activity owing to *SPTLC1* and *SPTLC2* variants and highlight potential decreases in circulating canonical sphingolipids that also may be relevant for HSAN1 pathophysiology.

**Table 2:** Incidence of *SPTLC1* and *SPTLC2* variants across HSAN1 patient cohort

| Variant | HSAN1 patients included | MacTel affected/predicted | MacTel unaffected |
| --- | --- | --- | --- |
| SPTLC1 C133Y | 8 | 8 | - |
| SPTLC1 C133W | 11 | - | 11 |
| SPTLC1 V144D | 1 | - | 1 |
| SPTLC2 A182P | 2 | - | 2 |
| SPTLC2 N177H | 1 | - | 1 |
| SPTLC2 S384F | 2 | 1 | 1 |
| **TOTAL** | **25** | **9** | **16** |

**Figure 4.**
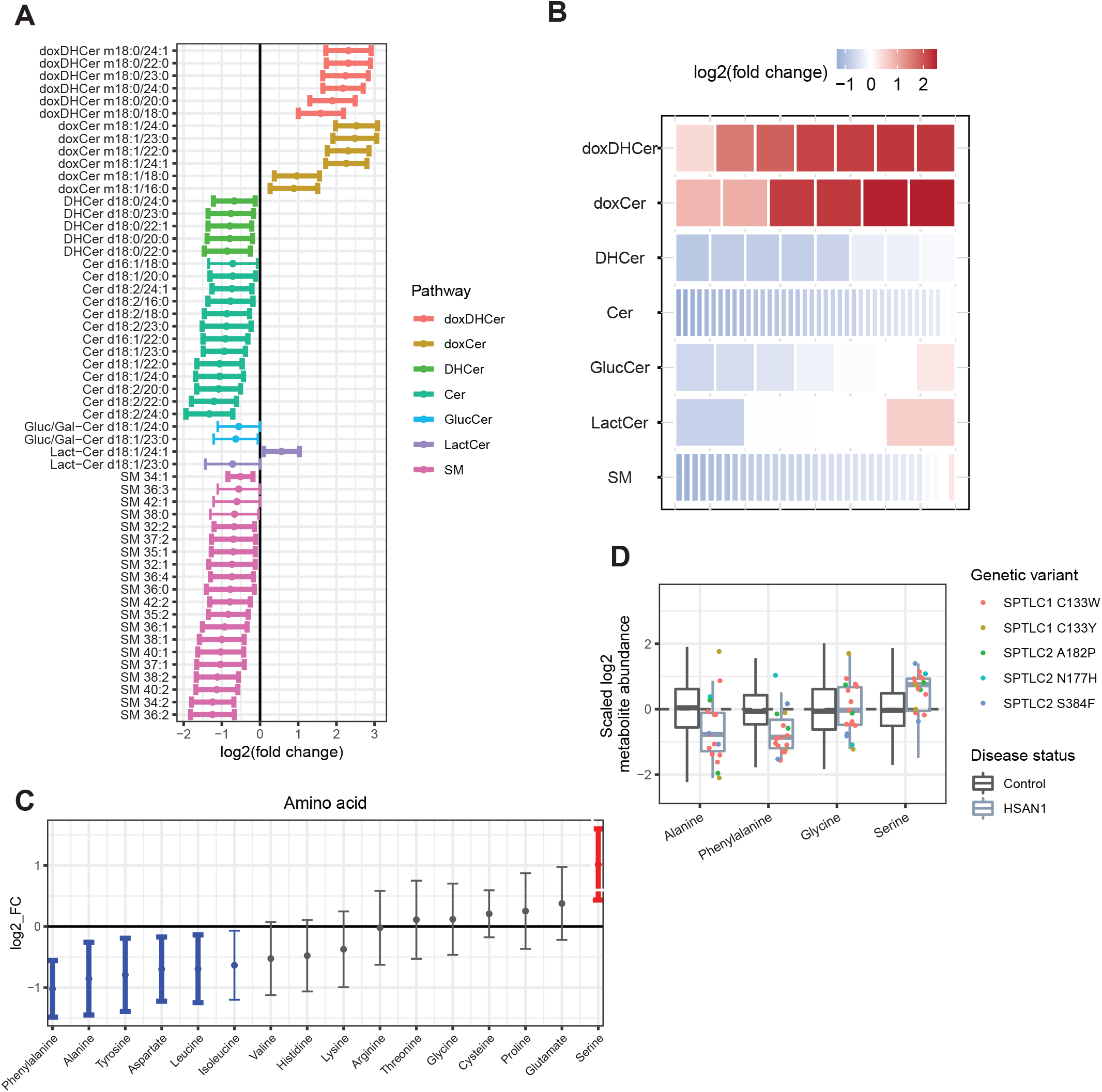
HSAN1 patients have altered circulating levels of canonical ceramides and amino acids A. Representation of all significantly different SL species (p<0.05) comparing HSAN1 to Controls. Color indicates the lipid class and bold bars indicate significance after correction for FDR. B. Changes in abundance across SL classes. Each row is composed of tiles representing lipids contained in the group. The color of each tile represents the mean log fold change of that lipid in HSAN1 patients to Controls. The color blue represents depletion and red represents increased abundance. C. Change in 16 amino acids comparing HSAN1 to Controls. Points indicate mean log fold change and bars extend to the 95% confidence interval of the mean. Red or blue colors indicate nominal significance (p<0.05), and bold bars indicate significance after correction for FDR. D. Distribution of alanine, phenylalanine, glycine, and serine levels in HSAN1 subjects (excluding those on serine supplementation) compared to Controls. HSAN1 subject metabolite abundance (points) are colored according to genetic variant. Individual data points for Controls are not shown. Boxplot central line indicates the median and box boundaries denote the 25th and 75th percentiles. Whiskers extend to the most extreme values within 1.5x the interquartile range. Abbreviations: doxDHCer, deoxydihydroceramide; doxCer, deoxyceramide; DHCer, dihydroceramide; Cer, ceramide; Gluc/Gal-Cer or GlucCer, glucosyl/galactosyl-ceramide; Lact-Cer, lactosyl-ceramide; SM, sphingomyelin; SPTLC1 and SPTLC2, serine-palmitoyl transferase long chain subunit 1 or 2

Intriguingly, HSAN1 patients exhibited higher serine levels and lower alanine levels compared to controls, showing the opposite effect as observed for MacTel patients (**Figure 4C**). These findings are not driven by a specific SPTLC variant and are observed with or without the serine supplemented patients (**Figure 4D, Supplemental Figure 7H**). Consequently, the correlations between serine or alanine and doxSLs were not present in these patients (**Supplemental Figure 2C**). HSAN1 patients also showed lower circulating phenylalanine, tyrosine, aspartate, and leucine concentrations, with a trend for reduced levels of the other BCAAs (**Figure 4C**). These amino acid changes suggest potential compensatory mechanisms to deal with the increased deoxysphingolipid levels (higher serine/lower alanine) and highlight distinctions in how these analytes might contribute to MacTel versus HSAN1.

### MacTel patients have distinct sphingolipid changes compared to patients with HSAN1

To determine if circulating levels of amino acid and sphingolipid can distinguish HSAN1 patients with or without MacTel (**Table 2**) we compared metabolite levels between these groups. This analysis identified several metabolites that were significantly different (uncorrected p<0.05) in HSAN1 patients +/-MacTel. These species were all lipids and included doxDHCer m18:0/24:1, doxCer m18:1/24:1, and Cer d18:1/24:1 that were less abundant in HSAN1 patients with MacTel (**Figure 5A; Supplemental Table 6**). Interestingly, these relatively abundant molecules all have a common n-acyl chain in nervonic acid (24:1). Other species that were lower in HSAN1 patients with MacTel patients included canonical SMs and lower abundant hexosylceramide (d18:1/20:0). This comparison suggests that HSAN1 patients with MacTel have more severe decreases in circulating canonical sphingolipids.

We next compared each patient subgroup (MacTel without SPT variants: MacTel only, n=205; HSAN1 without MacTel: HSAN1 only, n=16; HSAN1 with MacTel: MacTel + HSAN1, n=9) to control subjects, n=151. As noted above, HSAN1 patients (**Figure 4C**) had distinct amino acid changes compared to the MacTel group (**Figure 1A**) with serine increasing and alanine decreasing in HSAN1 patients, and these trends were maintained regardless of the MacTel status in the HSAN1 cohort (**Supplemental Fig 8A, Supplemental Table 8**). Although 1-deoxysphingolipids were high in all cohorts compared to controls, they were generally more elevated in HSAN1 only patients than MacTel + HSAN1, or MacTel only (**Figure 5B-C**). This finding is consistent with *in vitro* results indicating higher activity of SPTLC1^C133W^ versus the SPTLC1^C133Y^ variants^32-34^. MacTel + HSAN1 patients showed a broader trend for having lower circulating canonical ceramides and SMs compared to those patients with HSAN1 only (**Figure 5D, Supplemental Figure 8B-C**), and these patients had the lowest SM of all patient subgroups (**Figure 5E-F**), while no general trends were evident for the Lact-Cer and Gluc/Gal-Cer in these comparisons (**Supplemental Figure 8D-E**). Collectively, these results suggest that patients with HSAN1 and MacTel have more significant depletions in circulating canonical sphingolipids and highlight roles for both altered amino acids and membrane lipid homeostasis in driving MacTel.

**Figure 5.**
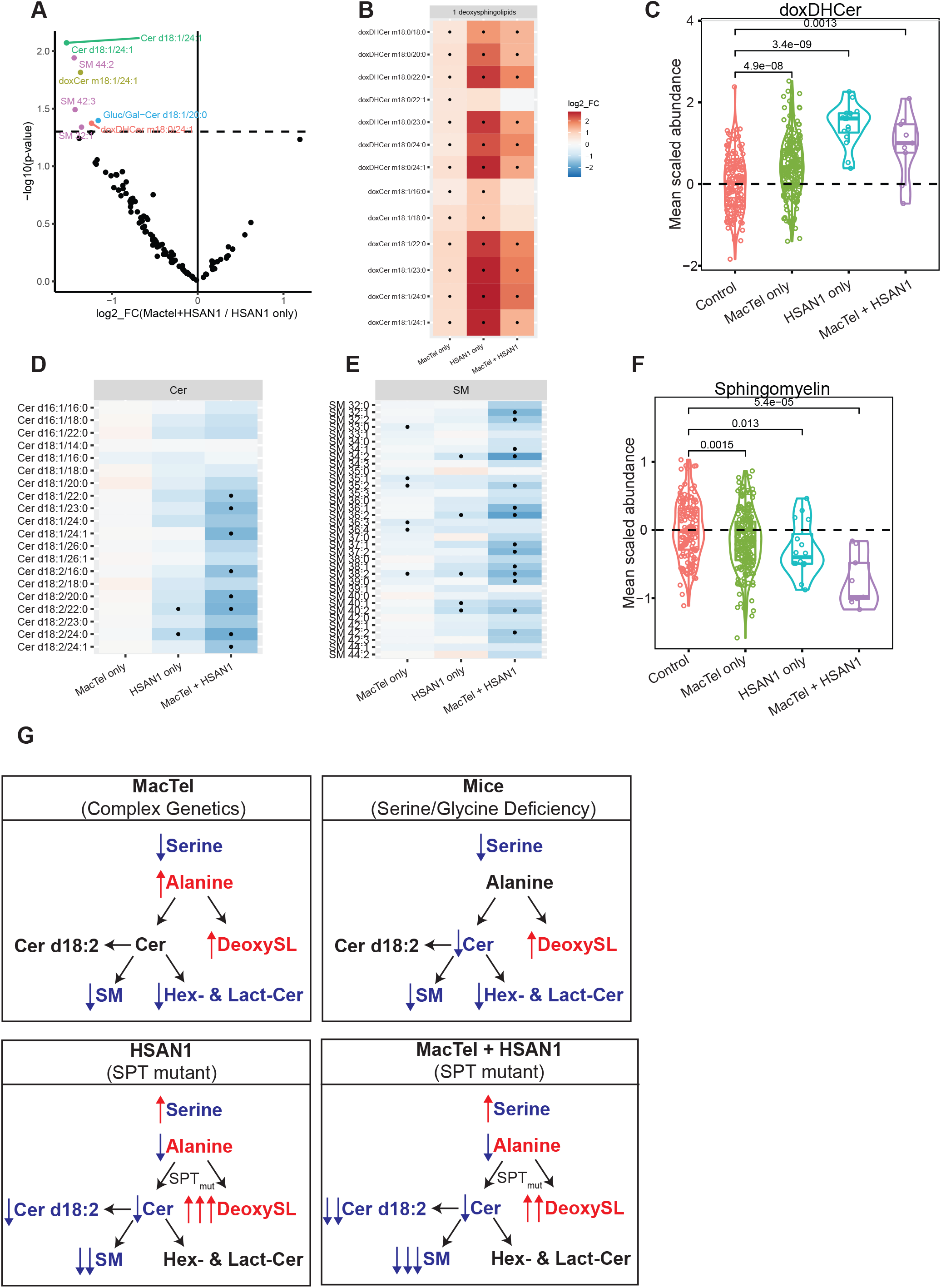
MacTel patients have distinct SL alterations compared to patients with HSAN1 A. Volcano plot comparing HSAN1 patients with MacTel, to HSAN1 patients without MacTel. Dashed line represents p=0.05. Only metabolites below this threshold are labeled and coloured by metabolite class. B. Depiction of the log2 fold change of 1-deoxysphingolipids in MacTel only, HSAN1 only, and HSAN1 patients with MacTel, compared to Controls. Tiles indicate the mean log fold change in abundance between each cohort (x) and controls. Black points indicate FDR-significant changes. Red colors indicate increased relative abundance, and blue colors indicate decreased abundance. C. Scaled abundance of averaged dihydroxyceramide levels in Controls, MacTel only, HSAN1 only, and HSAN1 patients with MacTel. Center line indicates the median and box limits indicate the 25th and 75th percentiles. D. Depiction of the log2 fold change of d18:2 ceramides in MacTel only, HSAN1 only, and HSAN1 patients with MacTel, compared to Controls. Tiles rendered as for panel B. E. Depiction of the log2 fold change of sphingomyelins in MacTel only, HSAN1 only, and HSAN1 patients with MacTel compared to Controls. Tiles rendered as for panel B. F. Scaled abundance of averaged sphingomyelin levels in Controls, MacTel only, HSAN1 only, and HSAN1 patients with MacTel. Center line indicates the median and box limits indicate the 25th and 75th percentiles. G. Summary of the amino acid and SL alterations in MacTel, HSAN1, and HSAN1 patients with MacTel compared to Controls. Red indicates increased, and blue indicates decreased abundance. Abbreviations: doxDHCer, deoxydihydroceramide; doxCer, deoxyceramide; DHCer, dihydroceramide; Cer, ceramide; Gluc/Gal-Cer or GlucCer, glucosyl/galactosyl-ceramide; Lact-Cer, lactosyl-ceramide; SM, sphingomyelin

## DISCUSSION

Here we comprehensively profiled serum amino acids and sphingolipid species in subgroups of patients with a rare polygenic macular disease (MacTel) and an even more rare autosomal dominant disease (HSAN1), two diseases that show clinical, metabolic, and genetic overlap. These metabolite groups intersect at an interesting biochemical node linking amino acid metabolism to membrane lipids important for nervous system function. In expanding the cohort and analytical complexity, we have confirmed metabolic changes in these patient populations (reduced serine/glycine and increased 1-deoxysphingolipids) and identified novel metabolite classes likely involved in disease pathology. The significant increases observed in alanine and glutamate levels in MacTel patients is further indication that amino acid dysregulation is a key contributor to MacTel. These amino acids become elevated in the context of mitochondrial dysfunction^35,36^ and highlight potential links between MacTel, deoxysphingolipids, and mitochondrial function^35,37-39^. Furthermore, we also find reduced serine/glycine and elevated BCAAs in MacTel patients versus control subjects, a pattern shared with obese and insulin-resistant patients^26^. MacTel patients have an increased prevalence of T2D^40^, which was reflected in our cohort and controlled for statistically. As such, these results provide additional evidence that MacTel and T2D share genetic drivers that impact amino acid metabolism through related mechanisms^5,41^.

In participants without SPT variants, we find that serine, glycine, and alanine strongly correlate with deoxydihydroceramide, but not the far more abundant canonical dihydroceramides. Consequently, monitoring deoxysphingolipid levels could function as a more stable and reliable biomarker of serine levels than the amino acid itself, which varies with diet and time of day^4^. This could be beneficial in both characterizing chronic serine levels as well as monitoring a patient’s response to treatment.

We observed distinct changes in canonical ceramides and complex sphingolipids in MacTel and HSAN1 patients. As summarized in Figure 5G, MacTel patients are typified by altered serine, glycine, and alanine levels and have elevated deoxysphingolipids and broad decreases in complex membrane sphingolipids but not ceramides. The HSAN1 patients, with altered SPT function, have elevated deoxysphingolipids, reduced ceramides and SMs, but the other complex sphingolipids were not generally affected. The mice with very low serine and glycine levels have elevated deoxysphingolipids and a reduction of all canonical sphingolipids, with ceramides and Gluc/Gal-Cers being the most reduced. The common feature between the three models is elevation of deoxysphingolipids and reduction in SM, suggesting that deoxysphingolipids might negatively influence SM levels. While no correlation was found between deoxysphingolipids and SM in the patient cohort, this mechanism fits with recent work by Clark *et al*, showing that HSAN1 patient cells had impaired membrane sphingolipid function^42^. Although the HSAN1 cohort is relatively small, those HSAN1 patients with MacTel had even lower levels of SM in circulation. On the other hand, we observed reduced Gluc/Gal-Cers and Lact-Cers specifically in MacTel patients and in the dietary mouse model, but not in HSAN1 patients. Since the amino acid levels are a distinguishing feature between the MacTel and HSAN1 patients, we speculate that low serine and glycine levels may be responsible for regulating these glycosylated complex sphingolipids. Furthermore, since the ceramides are not reduced in MacTel, it is likely via an SPT-independent mechanism.

Intriguingly, serine levels were generally elevated in HSAN1 patients, whereas canonical ceramides were broadly reduced compared to controls. In addition to dihydroceramide (d18:0) and ceramide (d18:1), the d18:2 ceramides were particularly impacted in the HSAN1 patients. This is interesting given that the enzyme responsible for their synthesis, FADS3, is also used in doxCer production^43^. The elevation of serine and low canonical ceramide levels suggest that sphingolipid supplementation (rather than serine) may be most beneficial therapeutically for these rare patients^10,33^.

These analyses indicate that while elevated deoxysphingolipids are likely integral to MacTel, they alone are not sufficient to drive the disease. Here we find that reduction in membrane sphingolipids might be an additional metabolic stressor. Indeed, our analysis of retinal imaging in MacTel patients indicated that low SM correlated with retinal neovascularization. Though no direct correlations have previously been described between neovascularization and SM, others have observed associations between ceramides and sphingosine 1-phosphate with ocular neovascularization and angiogenesis^44^. These results, combined with the reduction in glycosylated sphingolipids, further link MacTel pathogenesis to dysregulation of membrane lipids^45^. These data further support the concept that MacTel patients may benefit more directly from serine supplementation to improve amino acid and sphingolipid homeostasis compared to HSAN1 patients. However, further studies and more longitudinal data are needed to better understand the impact of systemic metabolic changes on the retinal phenotype and disease progression in MacTel.

## Data Availability

All data produced in the present study are available upon reasonable request to the authors.

## Supplemental Figure legends

**Supplemental Figure 1**. Effect of HSAN1 patient inclusion on metabolite expression

A. Comparison of the imputation results for the data containing all patient samples, compared with a subset of the data excluding HSAN1 patients

B. Scaled abundance of alanine and glutamate (left panel) and glycine and serine (right panel) in Control and MacTel. Center line indicates the median and box boundaries denote the 25th and 75th percentiles.

C.Correlation table of amino acids and lipids principal components in MacTel patients (left panel) and difference between correlation tables of Control and MacTel patients (right panel). Numbers indicate Pearson’s correlation coefficient (R).

**Supplemental Figure 2. MacTel patients have elevated doxSL levels and reduced SM**

A. Uncorrected abundance of deoxyceramides in Control and MacTel patients.

B. Uncorrected abundance of ceramides in Control and MacTel patients. Yellow box indicates the same species present as a deuterated internal standard.

C. Uncorrected abundance of glucosyl/galactosyl- and lactosylceramides in Control and MacTel patients.

D. Uncorrected abundance of sphingomyelin in Control and MacTel patients. Yellow box indicates the same species present as a deuterated internal standard. *nominally significant; **statistically significant with FDR<0.05

E. Association between serine (left) and alanine (right) abundance (x axes) and the average deoxydihydroceramide abundance (y axis) within each subject, colored according to patient cohort. Summary statistics are displayed for linear regression and colored according to patient cohort. R: Pearson’s correlation coefficient; p: p value.

**Supplemental Figure 3**. Abundance of SL species in mouse plasma on a serine/glycine-free diet

A. Uncorrected abundance of 1-deoxySL in mouse plasma on Control or serine/glycine-free (-SG) diet

B. Uncorrected abundance of ceramides in mouse plasma on Control or -SG diet

C. Uncorrected abundance of glucosyl/galactosyl- and lactosyl-ceramides in mouse plasma on Control or -SG diet

D. Uncorrected abundance of sphingomyelin in mouse plasma on Control or -SG diet. Yellow box indicates the same species present as a deuterated internal standard.

*nominally significant; **statistically significant with FDR<0.05

**Supplemental Figure 4**. Relationship between SM and hyper-reflectivity

Association between principal component value of the sphingomyelins metabolites across 100 MacTel patients. Both right eye (OD) and left eye (OS) are presented here as well as their sum (Union, OU). Hyper-reflectivity (A) and staging levels (B) defined by the presence of neovascularization (NV) are highlighted in the figure.

**Supplemental Figure 5**. Impact of excluding serine-supplemented subjects

A. Correlation of the effect of including serine-supplemented subjects with the effect of excluding serine-supplemented subjects.

**Supplemental Figure 6**. HSAN1 patients have elevated doxSL and lower levels of most other SL

A. Uncorrected abundance of 1-deoxySL in Control and HSAN1 patients.

B. Uncorrected abundance of ceramides in Control and HSAN1 patients. Yellow box indicates the same species present as a deuterated internal standard.

C. Uncorrected abundance of glucosyl/galactosyl- and lactosylceramides in Control and HSAN1 patients.

D.Uncorrected abundance of sphingomyelin in Control and HSAN1 patients. Yellow box indicates the same species present as a deuterated internal standard.

*nominally significant; **statistically significant with FDR<0.05

**Supplemental Figure 7**. Relative abundance of SL in HSAN1 patients stratified by genetic mutation

A. Abundance of deoxydihydroceramide in Control and HSAN1 patients with the indicated *SPTLC1* or *SPTLC2* mutations.

B. Abundance of deoxyceramide in Control and HSAN1 patients with the indicated *SPTLC1* or*SPTLC2* mutations.

C. Abundance of dihydroceramide in Control and HSAN1 patients with the indicated *SPTLC1* or*SPTLC2* mutations.

D. Abundance of ceramide in Control and HSAN1 patients with the indicated *SPTLC1* or *SPTLC2* mutations.

E. Abundance of glucosylceramide in Control and HSAN1 patients with the indicated *SPTLC1* or*SPTLC2* mutations.

F. Abundance of lactosylceramide in Control and HSAN1 patients with the indicated *SPTLC1* or*SPTLC2* mutations.

G. Abundance of sphingomyelin in Control and HSAN1 patients with the indicated *SPTLC1* or*SPTLC2* mutations.

H. Abundance of significantly altered amino acids in Control and HSAN1 patients with the indicated *SPTLC1* or *SPTLC2* mutation and **including** patients supplementing serine.

**Supplemental Figure 8**. MacTel patients have distinct SL changes compared to patients with HSAN1

A. Heat map of the fold change of amino acids between patients with MacTel only, HSAN1 only, or MacTel and HSAN1 to Control

B. Heat map of the fold change of dihydroceramides between patients with MacTel only, HSAN1 only, or MacTel and HSAN1 to Control

C. Heat map of the fold change of ceramides between patients with MacTel only, HSAN1 only, or MacTel and HSAN1 to Control

D. Heat map of the fold change of hexosylceramides between patients with MacTel only, HSAN1 only, or MacTel and HSAN1 to Control

E. Heat map of the fold change of lactosylceramides between patients with MacTel only, HSAN1 only, or MacTel and HSAN1 to Control

## Table Legends

**Supplemental Table 1**: LCMS/MS transitions and retention times used to identify sphingolipids

**Supplemental Table 2**: Formulation of amino acid-adjusted murine diets

**Supplemental Table 3**: Summary statistics for comparisons of metabolite abundance between each of three patient groups, and controls.

**Supplemental Table 4**: Molar quantitation of sphingolipid levels in patient cohorts. Data are group mean and 95% confidence interval of indicated patient groups

**Supplemental Table 5**: Summary statistics results for the association between metabolic pathway principal components and MacTel phenotype grading.

**Supplemental Table 6**: Summary statistics results for the association between all individual metabolites and MacTel phenotype grading.

**Supplemental Table 7:** Disease and variant status of HSAN1 cohort

**Supplemental Table 8**: Summary statistics for comparisons of metabolite abundance between MacTel patients presenting HSAN1, and patients presenting HSAN1 alone.

## Acknowledgements

We thank the Lowy family for their support of the MacTel Project, the Lowy Medical Research Institute. M.B. was supported by an Australian National Health and Medical Research Council (NHMRC) Investigator Grant (GNT 1195236). B.R.E.A. was supported by a NHMRC Early Career Fellowship (GNT 1157776). This work was also supported by the Victorian Government’s Operational Infrastructure Support Program and the NHMRC Independent Research Institute Infrastructure Support Scheme (IRIISS) and by an unrestricted departmental grant from Research to Prevent Blindness to the Moran Eye Center of the University of Utah. This work was also supported by NIH grant R01CA234245 and NSF CAREER Award #1454425 (to C.M.M.)

Finally, the authors wish to thank the MacTel patients, HSAN1 patients and their families for their involvement in this study, and the Lowy Medical Research Institute for supporting this work.

## Author contributions

C.R.G.: conceptualization, formal analysis, data curation, investigation, methodology, writing—original draft, writing—review and editing, and visualization; R.B.: conceptualization, software, formal analysis, data curation, investigation, writing—original draft, writing—review and editing, and visualization; B.R.E.A.: conceptualization, software, formal analysis, data curation, investigation, writing—original draft, writing—review & editing, supervision project administration; S.T.: formal analysis, investigation, writing-original draft; M.H.: investigation, writing—review and editing; G.M.: investigation, writing— review and editing; B.H.: investigation; J.T.: investigation; M.M.R.: investigation; P.S.B.: investigation; writing—review and editing, C.E.: investigation; M.Fru.: investigation; M.Fri.: resources, writing-review & editing, supervision project administration and supervision; M.W: conceptualization, methodology, writing-review & editing; M.B.: review & editing, supervision project administration and supervision; C.M.M.: conceptualization, resources, supervision project administration and supervision, writing - review & editing; M.L.G.: conceptualization, data curation, formal analysis, investigation, project administration, resources, supervision, visualization, writing - original draft, review & editing.

