## Supplemental Figures for "Divergent amino acid and sphingolipid metabolism in patients with inherited neuro-retinal disease"

### Supplementary Figure 1

#### Amino acid levels in MacTel patients and impact of inclusion of HSAN1 patients

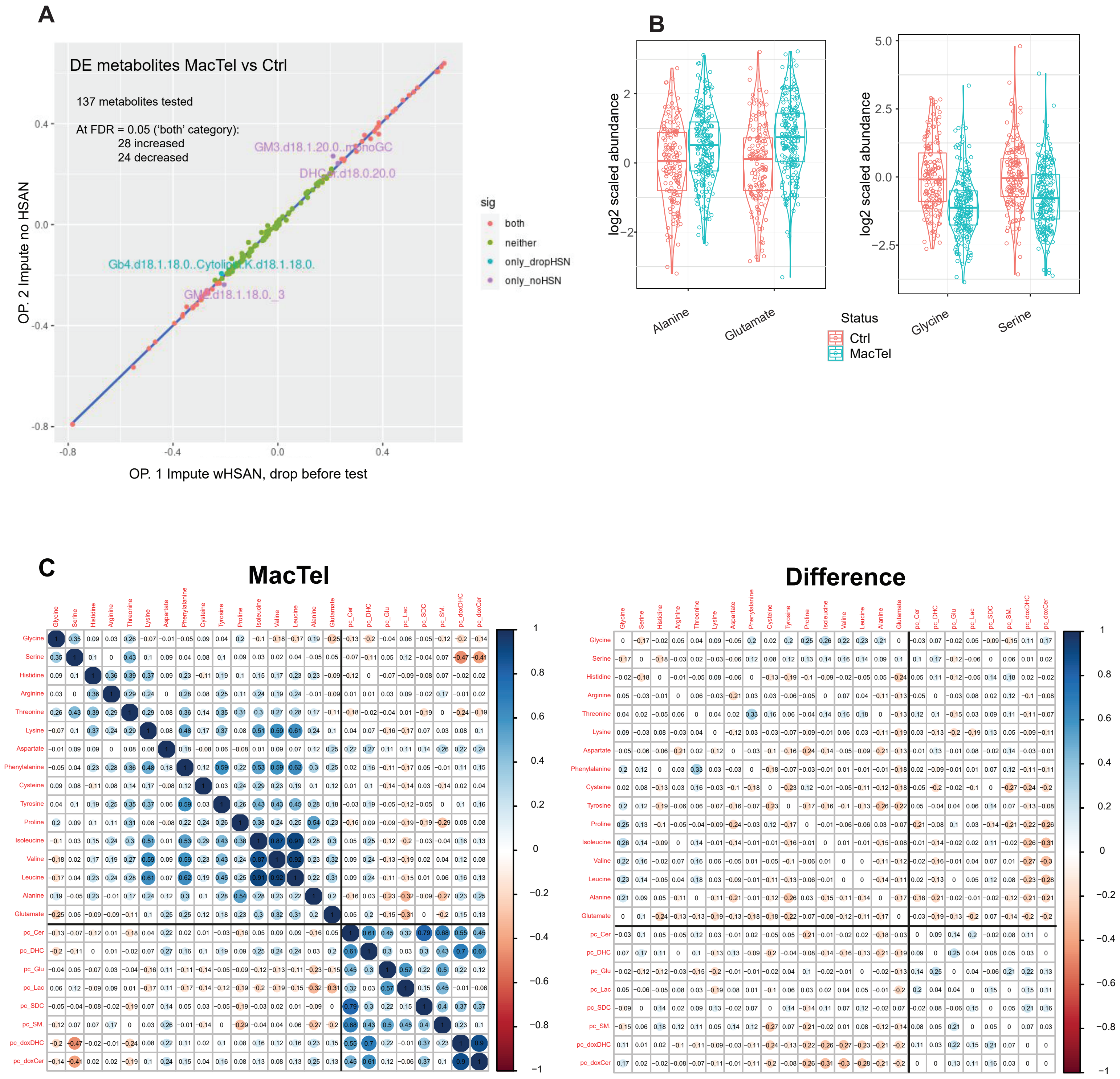

### Supplementary Figure 2

#### MacTel patients have elevated doxSL levels and reduced SM

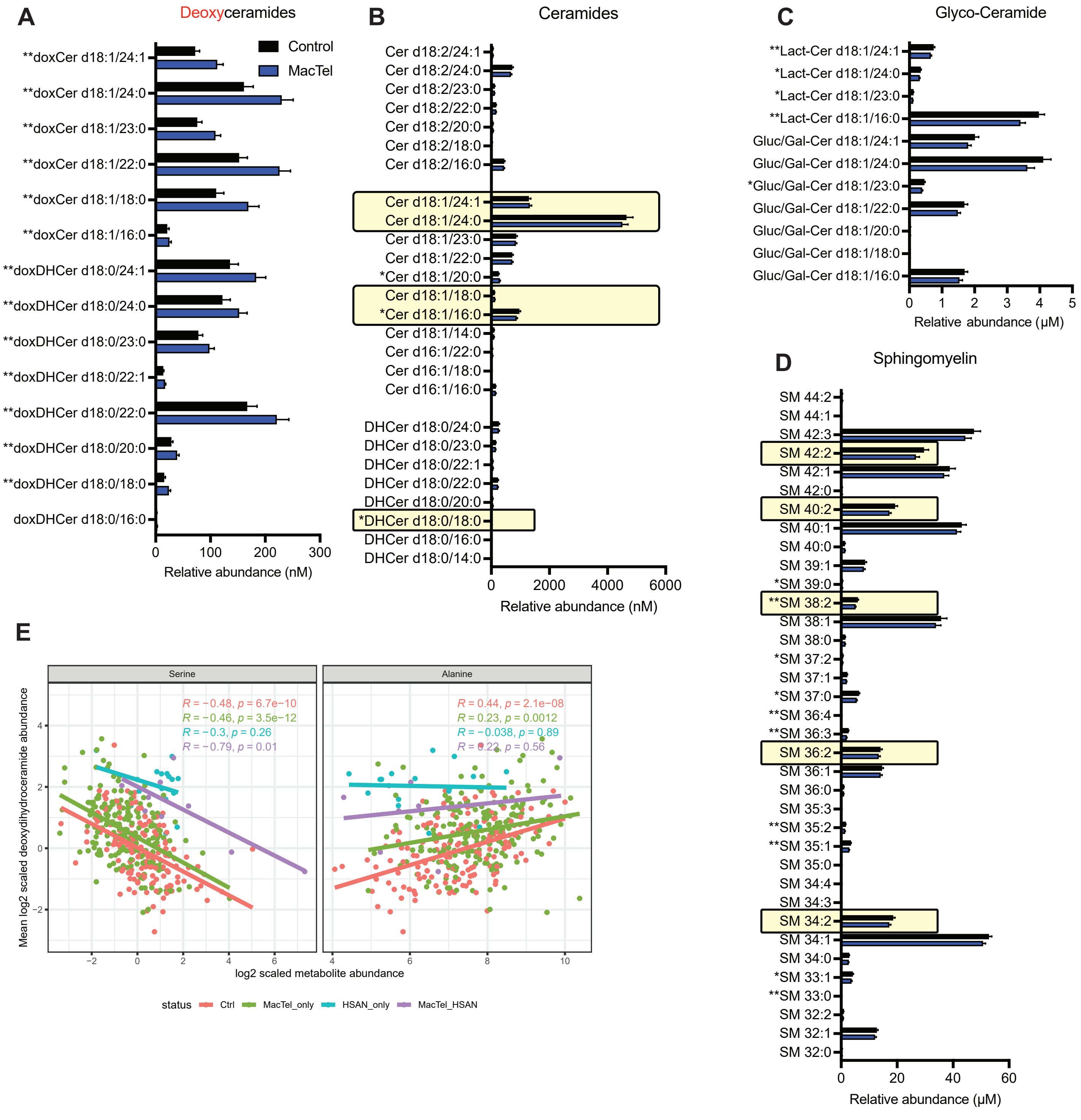

### Supplementary Figure 3
#### Abundance of SL species in mouse plasma on a serine/glycine-free diet

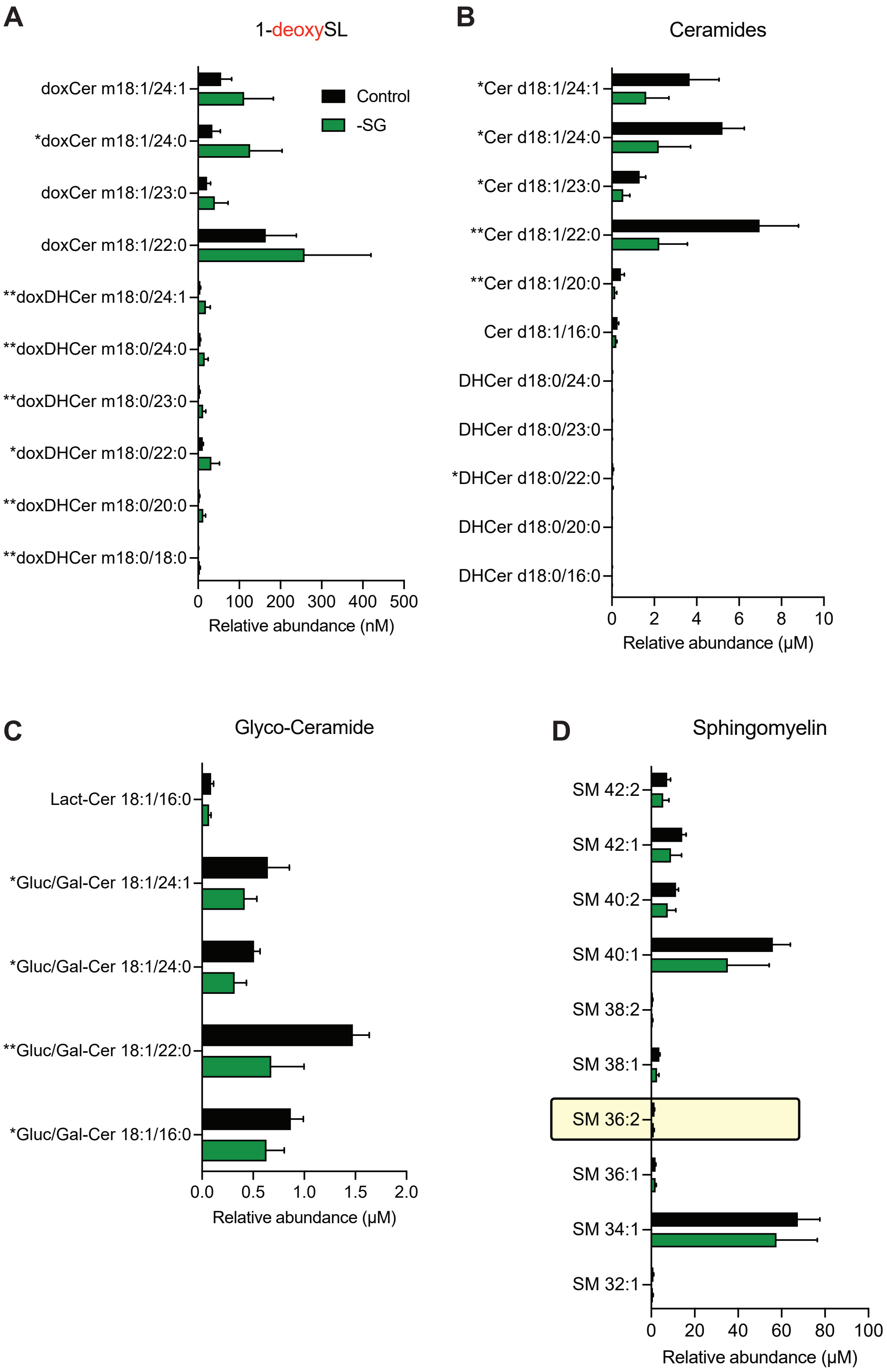

### Supplementary Figure 4

#### Relationship between SM and hyperreflectivity

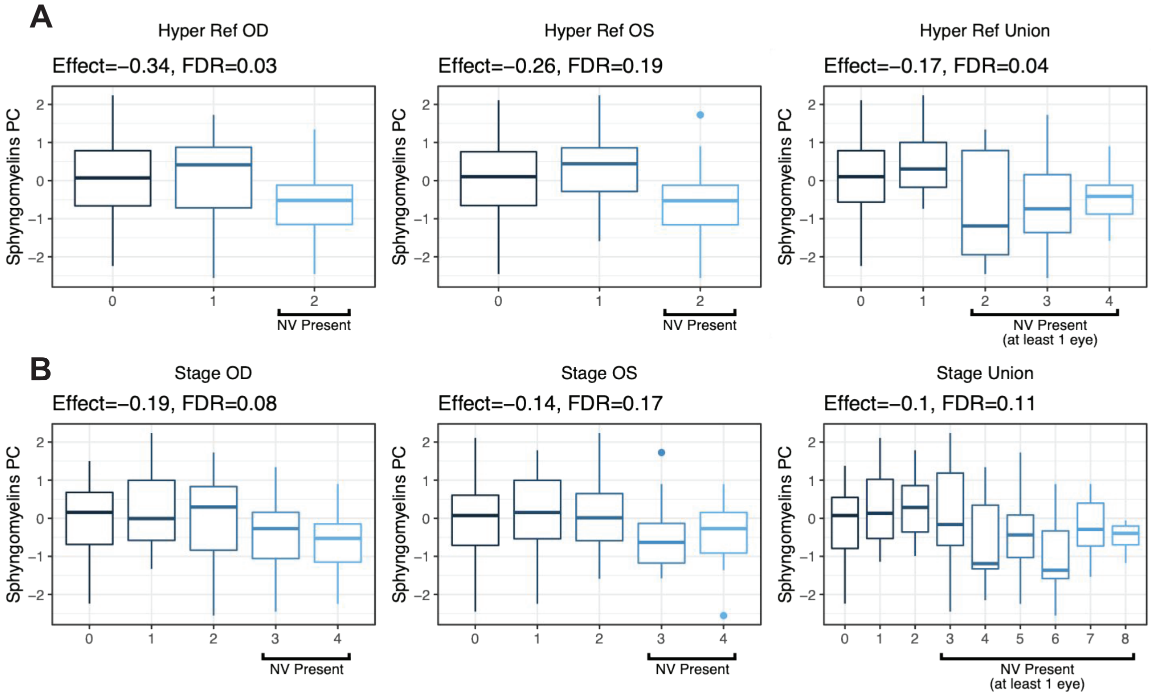

### Supplementary Figure 5

#### Impact of excluding serine-supplemented subjects

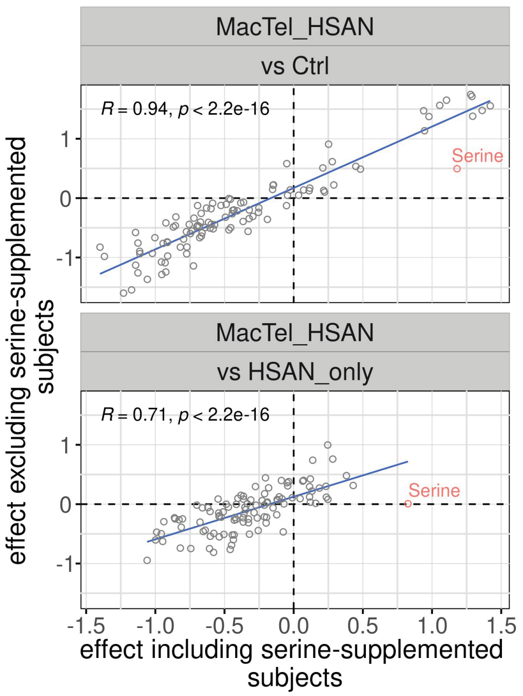

### Supplementary Figure 6

#### HSAN1 patients have elevated doxSL and lower levels of most other SL

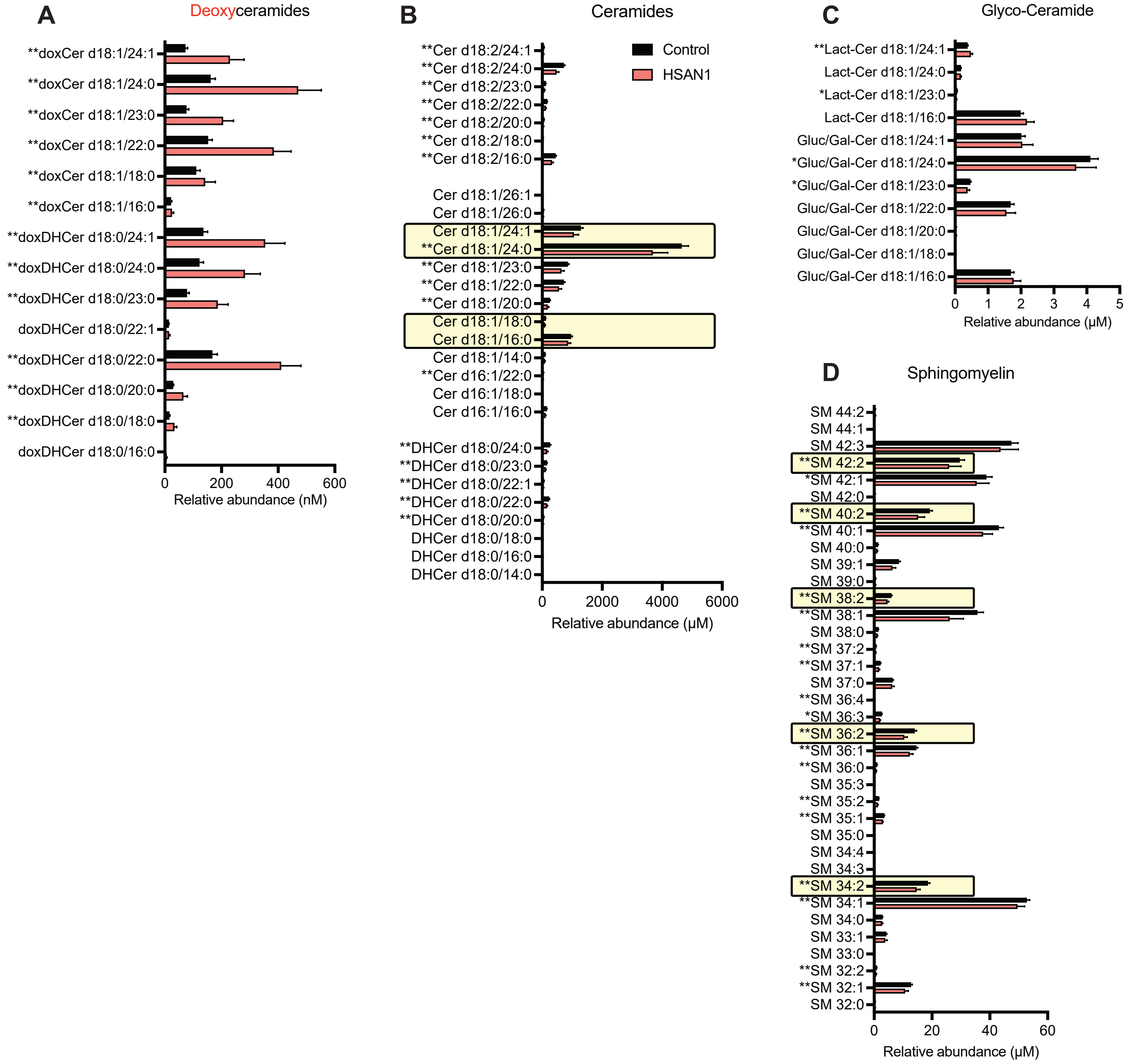

### Supplementary Figure 7
#### Relative abundance of SL in HSAN1 patients stratified by genetic mutation

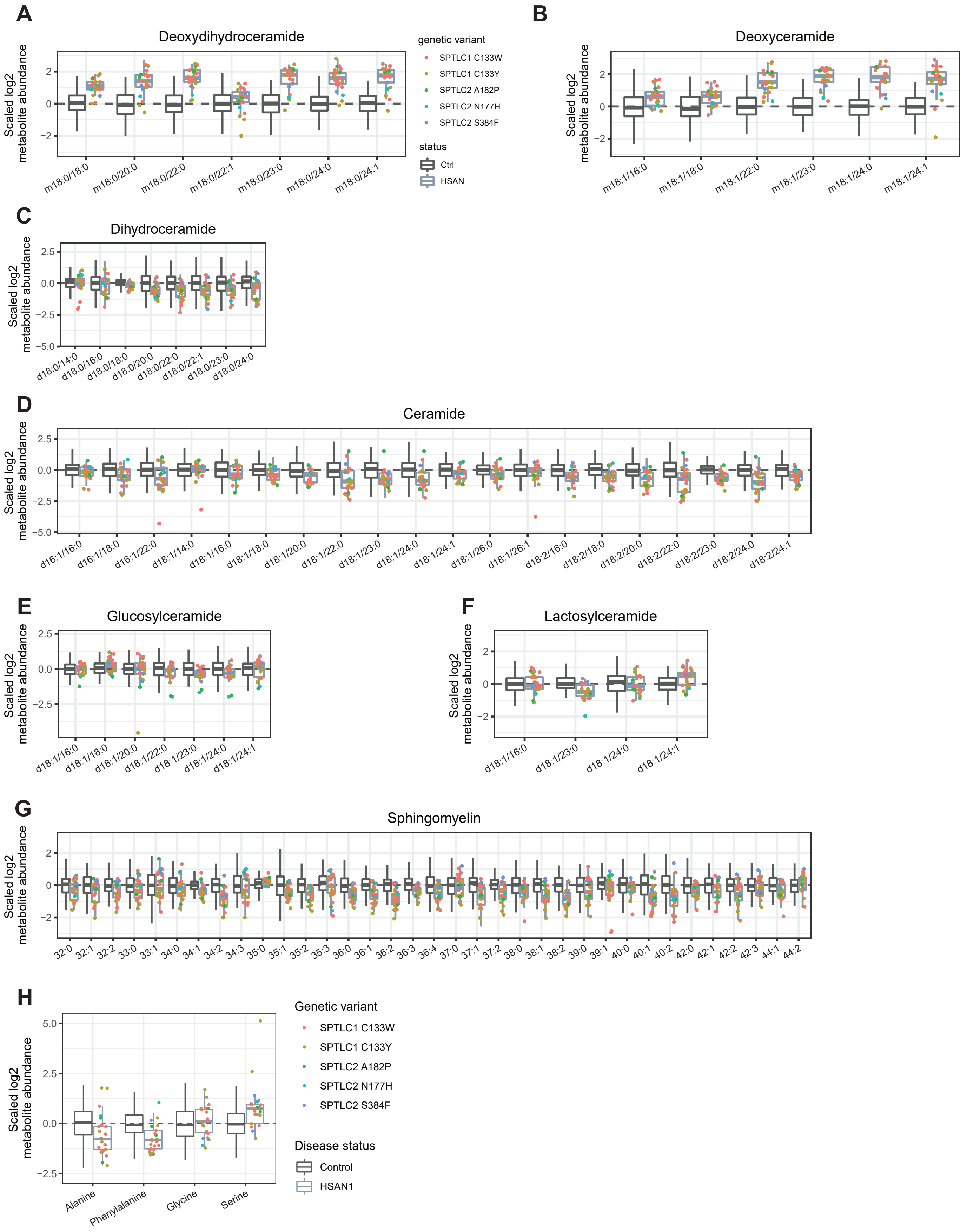

### Supplementary Figure 8

#### MacTel patients have distinct SL changes compared to patients with HSAN1

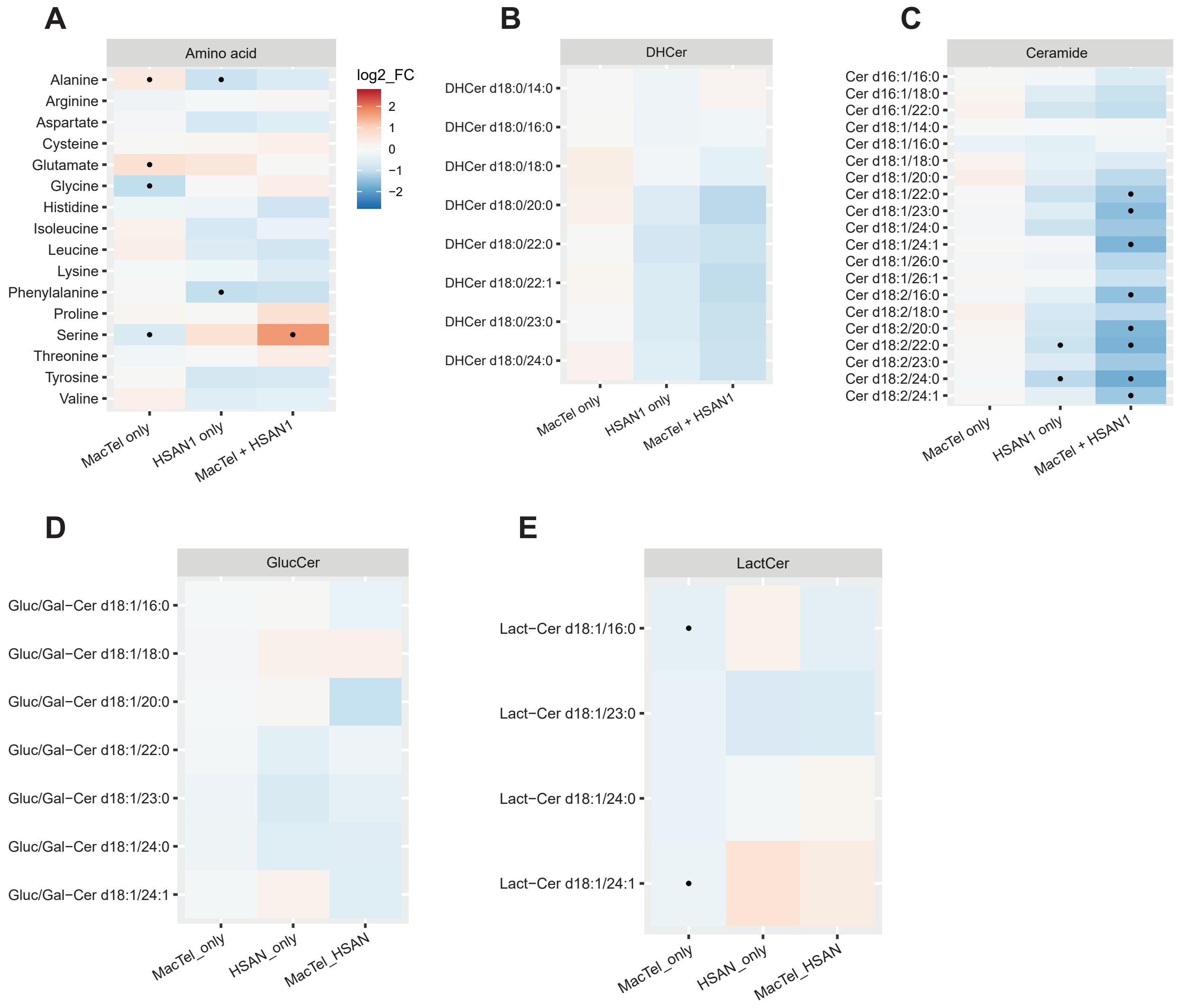
